# Deep transcriptome profiling of multiple myeloma with quantitative measures using the SPECTRA approach

**DOI:** 10.1101/2020.10.06.20206714

**Authors:** Rosalie Griffin Waller, Heidi A. Hanson, Brian J. Avery, Michael J. Madsen, Douglas W. Sborov, Nicola J. Camp

**Affiliations:** Huntsman Cancer Institute and School of Medicine, University of Utah, Salt Lake City, UT, 84112, USA; Computational Biology, Quantitative Health Sciences, Mayo Clinic, Rochester, MN, 55905, USA

## Abstract

SPECTRA is a new data framework to describe variation in a transcriptome as a set of unsupervised quantitative variables. Spectra variables provide a deep dive into the transcriptome, representing both large and small sources of variance, and are ideal for modeling alongside other variables for any outcome of interest. Each spectrum can also be considered a phenotypic trait, providing new avenues for disease characterization or to explore disease risk. We applied the SPECTRA approach to multiple myeloma (MM), the second most common blood cancer. Using RNA sequencing from malignant CD138+ cells, we derived 39 spectra in 767 patients from the MMRF CoMMpass study. We included spectra in prediction models for clinical endpoints, compared to established expression-based risk scores, and used descriptive modeling to identify associations with patient characteristics. Spectra-based risk scores added predictive value beyond established clinical risk factors and other expression-based risk scores for overall survival, progression-free survival, and time to first-line treatment failure. Significant spectra in models may provide mechanistic insight via gene set enrichment based on their gene weights. Gene set enrichment in CD138+ spectrum S5, which was significant for all prognostic endpoints, indicated enrichment for genes in the unfolded protein response, a mechanism targeted by proteasome inhibitors, common first line agents in MM treatment. We also identified significant associations between CD138+ spectra and tumor cytogenetics, race, gender, and age at diagnosis. The SPECTRA approach provides measures of transcriptome variation to deeply profile tumors with greater flexibility to model clinical outcomes and characteristics.

**AUTHOR SUMMARY:** Complex diseases, including cancer, are highly heterogeneous, and large molecular datasets are increasingly part of describing an individual’s unique experience. Gene expression is particularly attractive because it captures genetic, epigenetic, and environmental consequences. Transcriptome studies are gaining momentum in genomic epidemiology, and the need to incorporate these data in multivariable models alongside other risk factors brings demands for new approaches. The SPECTRA approach is a new intrinsic quantitative data framework for transcriptomes. A tissue is described by a set of quantitative measures (or ‘spectra’ variables) to deeply profile gene expression in a tissue. Spectra variables are independent and offer flexibility for use in predictive or descriptive modeling. We applied the SPECTRA approach to multiple myeloma, the second most common blood cancer. A set of 39 spectra variables were derived to represent the myeloma tumors. Outcome modeling provided SPECTRA-based risk scores that added predictive value for clinical outcomes beyond established risk factors.

## INTRODUCTION

Numerous factors are involved in risk and prognosis in complex disease. Transcriptomes represent the combined effects of inherited, somatic, and epigenetic variation and can provide insight into genetic and environmental risk factors. As a result, gene expression studies are gaining momentum in genomic epidemiology (Allott et al., 2020; López et al., 2019; Stopsack et al., 2018; Sweeney et al., 2014; Zhang et al., 2018). The need to incorporate transcriptome data in multivariable models alongside other risk factors brings new demands for approaches to describe transcriptomes.

Many current transcriptome approaches are focused on immediate biological interpretation and constrained to biological expectations or reduce the data to a single categorical variable (e.g., intrinsic subtypes). While these have advanced knowledge of disease mechanisms (Brunet et al., 2004; Tamayo et al., 1999; Way et al., 2020) and identified important high-level differences in disease (Lapointe et al., 2004; Perou et al., 2000; Shaughnessy et al., 2007), complementary approaches are needed to go deeper and increase flexibility and application. In common disease, the sources of heterogeneity are many, complex, and often poorly understood. Latent variables, focused on capturing signal and not immediate interpretability, provide the potential for new discoveries. Transcriptome characterization using multiple quantitative variables may provide a meaningful deeper dive into the transcriptome.

Here we describe the SPECTRA approach, a data workflow and variable derivation method with principal component analysis (PCA) at its core. PCA has many strengths aligned with our intent, such as providing multiple unsupervised variables that optimize coverage of the global variance. The resulting latent variables are quantitative, uncorrelated, retain integrity to the original data, and have desirable attributes for subsequent multivariable prediction and descriptive modeling. A limitation of PCA is that if data are not curated adequately, spurious variance will be incorporated in the derived variables, including technical artifacts and missing data structures. A key part of the SPECTRA approach is stringent data culling, quality control, zero-handling, and normalization.

For well-curated gene-panel data a PCA-based approach has shown promise. Using a population-based dataset of breast tumors, we previously used PCA to reduce the 50-gene space of the PAM50 gene panel to five quantitative multi-gene expression variables (Madsen et al., 2018). When implemented as predictor variables in an independent clinical trial dataset, PCA variables were able to predict prognosis and response to paclitaxel; adding value beyond clinical risk factors and outperforming intrinsic subtypes (Camp et al., 2019). When utilized as quantitative tumor phenotypes, PCA variables were superior to the standard PAM50 subtypes for gene mapping (Hanson et al., 2020; Madsen et al., 2018). Here, we describe a method to derive a quantitative data framework for whole transcriptomes.

**Figure 1** uses a color analogy to illustrate the conceptual framework of SPECTRA and contrast our goal of quantitative variables for direct use in outcome modeling with a more conventional categorization approach using hierarchical clustering. Each spectrum in **Figure 1** (𝑥*_R_*, 𝑥*_G_*, 𝑥*_B_*) are independent variables that can be directly used to model any outcome (*y_i_*), and other covariates/predictors can also be easily included (**Figure 1d**). Conversely, unsupervised hierarchical clustering uses the spectra to categorize patients into groups (**Figure 1c**), flattening the multiple variables to a single categorical variable which may reduce statistical power. For example, in **Figure 1**, *x_R_* cannot be represented by any group ordering in **Figure 1c**, and associations for that spectra variable would be lost. An alternate convention is to supervise clustering to an outcome. But, while supervised clustering can improve power over unsupervised clustering for the prediction of a single outcome, it also tethers the groups to the trained outcome and doesn’t facilitate comparison to other outcomes.

**Figure 1.**
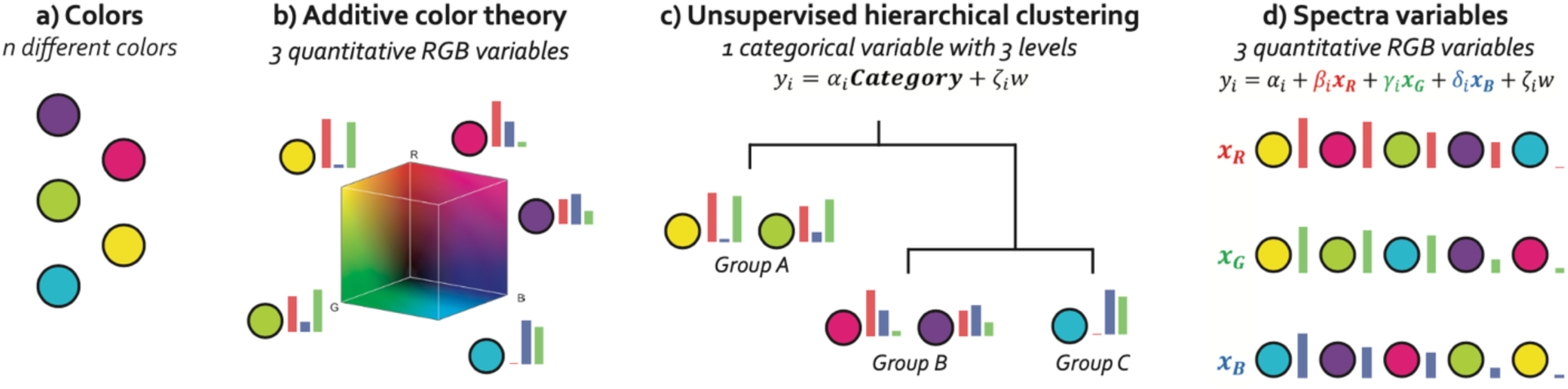
A color analogy to illustrate the advantages of spectra variables for modeling. a) <u>Individual observations</u> of color. b) <u>Dimension Reduction</u> (additive color theory), all colors can be represented using 3 quantitative RGB variables. c) <u>Standard-use</u>, modeling on the 3 RGB variables used to identify structure across samples using hierarchical clustering. This derives groups based on the complete 3-variable RGB profile to derive one polychotomous meta-variable (different groups are non-ordinal levels). d) <u>Multivariable modeling implementation of spectra variables</u>, multiple separate spectra integrated directly into a multivariable analysis. Each uncorrelated variable can be assessed separately for its predictive value for an outcome. This implementation retains the full resolution of the initial data because the variables are quantitative and retain integrity to the initial data. Note, lower-resolution versions of *x_B_* and *x_G_* can be achieved using hierarchical groups but the loss of quantification will likely also lose power. *x_R_* cannot be captured by any group ordering and associations for this spectrum would be lost using hierarchical groups.

We illustrate our whole transcriptome SPECTRA approach using bulk RNA sequencing (RNAseq) data from CD138+ sorted myeloma cells from the Multiple Myeloma Research Foundation (MMRF) CoMMpass Study (Keats et al., 2013). Multiple myeloma (MM) is a malignancy of the plasma cells with one of the poorest 5-year survival (55.6%) for adult-onset hematological malignancies (SEER, 2021). It is most frequently diagnosed at ages 65-74 years (median 69 years) (SEER, 2021). Incidence is higher in men (8.7 men vs. 5.6 women per 100,000) and particularly high in patients self-reporting as African American (AA men 16.3, and AA women 11.9 per 100,000) (SEER, 2021). We use the SPECTRA approach to derive quantitative CD138+ transcriptome variables, referred to as *spectra*, which we use in regression models to predict clinical endpoints, derive latent risk groups that may be clinically meaningful, compare to established expression-based molecular risk scores, and describe associations with clinical and demographic characteristics.

## RESULTS

### SPECTRA: an approach for deep transcriptome profiling with quantitative measures

The motivation is the derivation of well-behaved, quantitative variables from RNAseq data to capture transcriptome variation that can be used universally as predictors for any outcome, and as novel phenotypes. The approach requires an RNAseq dataset to derive the framework of transformations for the SPECTRA variables and multiple spectra are calculated for each individual in the dataset. An overview of the SPECTRA approach is shown in **Figure 2** and **Figure S1**. As an agnostic technique, the goal is to retain only those aspects of the RNAseq data that can represent meaningful variance. Accordingly, genes likely to lack precision are removed and only coding genes with sufficient coverage across the dataset are considered. An internal normalization procedure accounts for feature-length, library size, and RNA composition. This normalization avoids the need for reference samples, real or synthetic, and provides the potential for spectra to be ported to follow-up samples and external datasets. Finally, skew and outliers are dealt with before PCA is performed. Specific details are listed in Materials and Methods.

**Figure 2.**
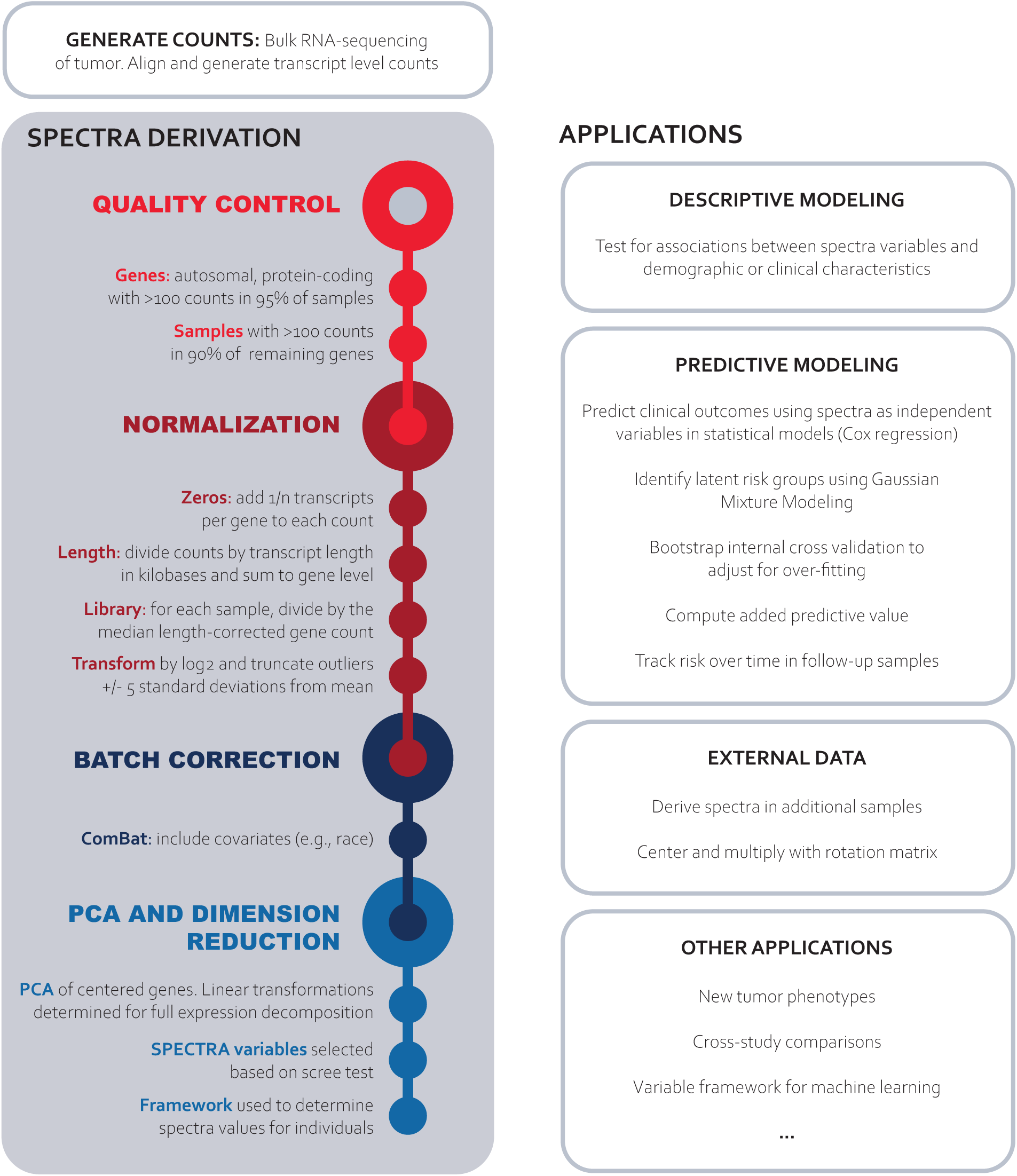
Overview of SPECTRA framework to derive spectra variables and example applications using spectra variables.

PCA is a well-established, unsupervised, data-driven method that, based on the covariance of a dataset, produces a matrix factorization which is a unique solution of linear transformations and transformed values. The linear transformations preserve the variance in the data and provide meaningful comparisons between individuals. The transformed values are orthogonal quantitative variables (linearly uncorrelated), each subsequently going deeper into the global variance. Dimension reduction is achieved by selecting only the first *k* components, for which the proportion of total variance explained can be described. Details of the matrix factorization performed by PCA can be found in Supplemental Methods.

The results of the subsequent PCA are the rotation matrix that describes the multi-gene linear transformations; and the transformed data matrix, the quantitative variables for each individual referred to as *<u>SPECTRA variables</u>*, or *<u>spectra</u>*. The set of linear transformations provides a new reduced-dimension *<u>framework</u>* for the expression space. The SPECTRA variables are linearly independent, each providing additional coverage of the variance, and as unsupervised variables can be used as predictors for any outcome and as novel phenotypes.

### CD138+ Spectra

We applied the SPECTRA approach to RNAseq from treatment naïve CD138+ cells collected at diagnosis from 768 patients in the MMRF CoMMpass study (Keats et al., 2013). We used transcript-based expression estimates generated by the CoMMpass study with Salmon (Patro et al., 2017). After quality control, gene expression values for 7,449 genes (56,339 transcripts) in 767 patients were normalized and batch corrected. After PCA, the first 39 components were selected based on a scree test and standardized to create spectra S1-S39. Together these explain 65% of the global expression variation (**Figure 3A**). As linearly uncorrelated variables each of the 39 CD138+ spectra capture a different source of variance – each spectrum has the potential to describe patient differences. A patient’s CD138+ transcriptome spectra variables can be visualized with a 39-spectra barcode (**Figure 3B**).

**Figure 3.**
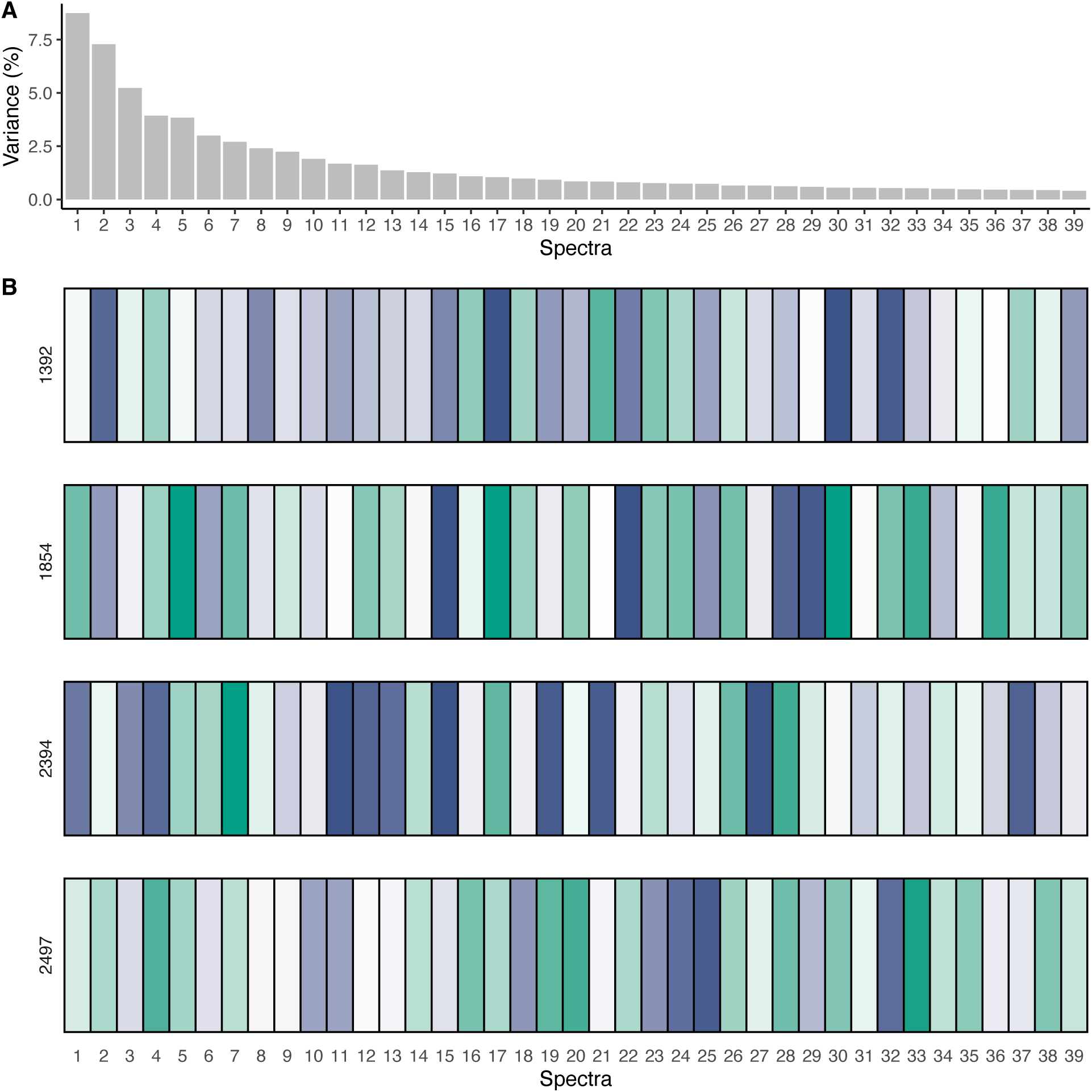
Spectra barcodes in four CoMMpass patients. A) Percent of transcriptome-wide variance between samples captured by each spectrum. Together the spectra capture 65% of the transcriptome-wide variance. B) For each patient, all 39 spectra are illustrated with the value represented by the bar intensity. The color indicates if the patient’s spectra value is positive (blue) or negative (green). Each patient has a unique profile across the 39 spectra. At a high-level patients 2497 & 1854 may appear most similar (mostly green) and 2394 & 1392 (mostly blue). However, at a finer resolution, similarities vary. For example, for spectrum S1 and S2, patients 2497 and 1854 are quite different. For spectrum S1, 2497 is more like 1392, for spectrum S15, 1854 is more like 2394.

### Predictive Modeling – Overall Survival (OS)

Predictive Modeling used Cox proportional hazards regression with bootstrap internal validation to adjust for over fitting (Harrell, 2015, pp. 114–116). This strategy maintains the full sample for discovery (referred to as ‘apparent’ results) and implements bootstrapping to estimate over-fitting (referred to as ‘optimism’). The optimism corrected apparent results are referred to as ‘adjusted’ values. In Cox regression of OS (179 events), nine spectra were significant (p < 0.05) and selected for the predictive model (apparent C-index = 0.70). Internal validation (1000 bootstraps) estimated C-optimism = 0.04, leading to an adjusted C-index (C_adj_ = 0.66 (0.65-0.74). Model coefficients are given in **Table 1**. Gaussian mixture modeling on the OS spectra risk scores identified two risk groups (p = 0.001) (**Figure 4**) with 88 patients in the high-risk group (58 events, median OS 20.1 months) and 676 patients in the low-risk group (121 events, median OS not reached after 78 months) (**Figure 4C**). After internal validation, the optimism adjusted hazard ratio (HR_adj_) for the high-compared to low-risk patients was 4.27 (2.31-12.69) (**Table 2**). This compared to UAMS OS HR_adj_ = 3.89 (2.53-5.45) and SBUK OS HR_adj_ = 2.54 (1.91-3.47) in CoMMpass data.

**Figure 4.**
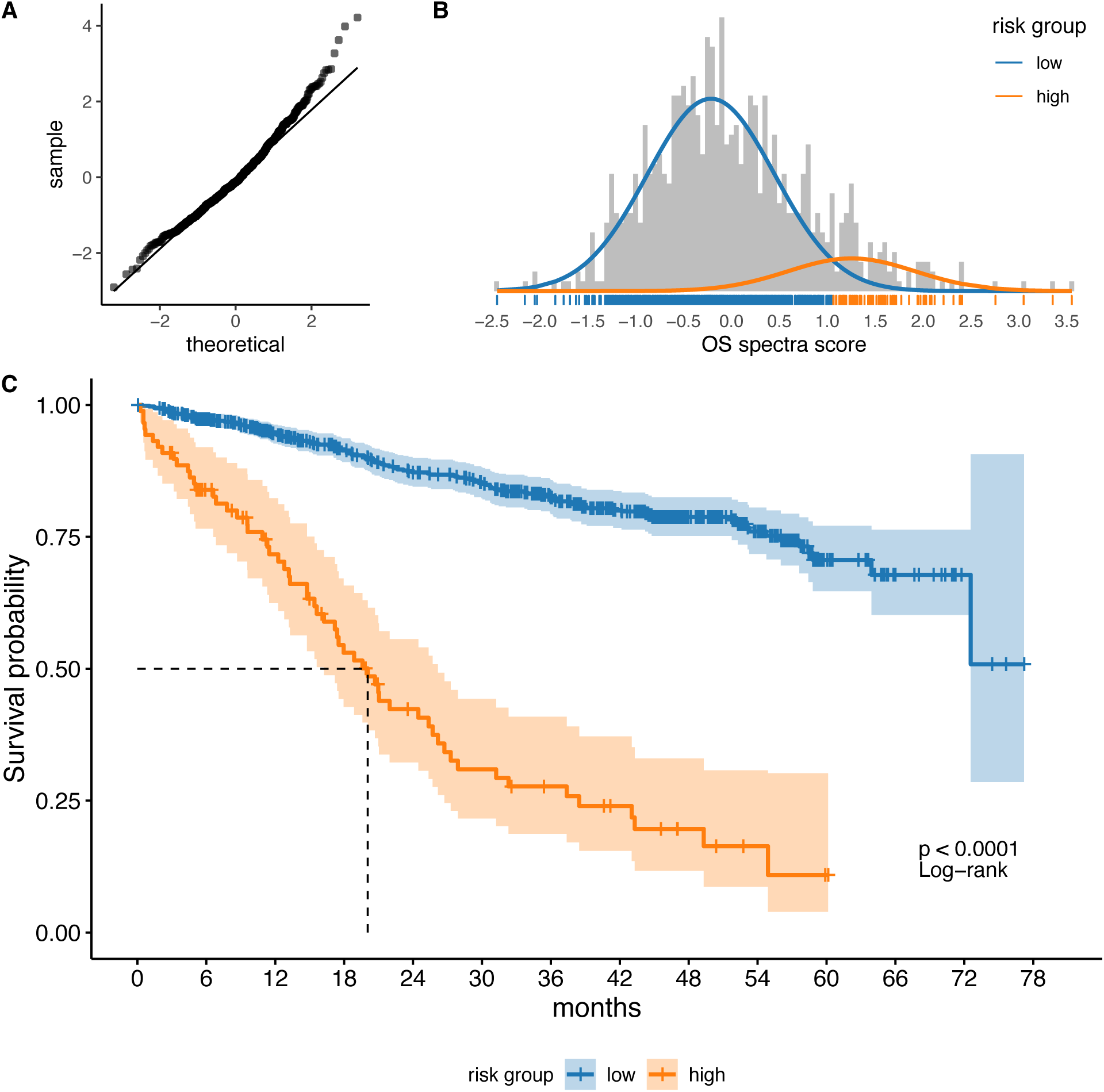
Overall survival spectra predictive modeling. A) Quantile-quantile plot of the actual spectra overall survival risk score (Cox linear predictors) compared to theoretical values. Elevated tail suggests high-risk group exists. B) Gaussian mixture modeling identified two distributions in the spectra score. Each patient was assigned into an OS risk group based on distribution probability. The blue bars on the x-axis represent patients in the low-risk group and the orange bars are patients in the high-risk group. C) Kaplan-Meier curve of the high/low risk groups. High-risk patients (n = 88, 58 events) had median survival of 20.1 months. Low-risk patients (n = 679, 121 events) did not reach median survival in the study timeframe.

**Table 1.** Cox regression beta coefficients for overall survival (OS), progression free survival (PFS), and time to treatment failure (TTF). Beta coefficients are by per spectra standard deviation. The quantitative spectra risk score for an outcome is the weighted sum of the spectra retained in its model based on their beta coefficients: ∑*_j_ β_j_S_j_*.

| Spectra | OS | PFS | TTF |
| --- | --- | --- | --- |
| 1 |  | -0.132 | -0.218 |
| 2 | -0.335 | -0.241 | -0.212 |
| 3 | 0.243 |  |  |
| 4 | 0.461 | 0.282 | 0.182 |
| 5 | -0.324 | -0.271 | -0.257 |
| 9 | -0.335 | -0.182 | -0.176 |
| 10 |  |  | -0.138 |
| 11 |  | 0.083 |  |
| 12 | -0.179 |  |  |
| 13 | 0.174 |  |  |
| 18 |  |  | 0.131 |
| 19 | 0.111 |  |  |
| 24 |  | 0.178 | 0.118 |
| 26 | 0.184 | 0.101 |  |
| 31 |  |  | 0.102 |
| 32 |  | 0.121 | 0.117 |

**Table 2.** Overall survival hazard ratios between high-and low-risk patients using spectra, UAMS, or SBUK to define patient groups.

| **Risk Score** | **Apparent HR** | **Adjusted HR** | **Optimism** |
| --- | --- | --- | --- |
| OS Spectra | 6.44 (4.47-14.86) | 4.27 (2.31-12.69) | 2.16 |
| UAMS | 3.98 (2.62-5.54) | 3.89 (2.53-5.45) | 0.09 |
| SBUK | 2.58 (1.95-3.51) | 2.54 (1.91-3.47) | 0.04 |

The added predictive value (APV) of the quantitative spectra risk score, beyond Revised International Staging System (R-ISS) and age at diagnosis was 0.612 (**Table 3**). In parallel analyses, UAMS APV was 0.576 and SBUK APV = 0.502, indicating each contains substantial predictive value beyond clinical factors. Compared to a model including R-ISS, age at diagnosis, and UAMS, however, SBUK showed very little added value (APV=0.006, **Table 3**), indicating its predictive gene expression information is almost all duplicative with UAMS. In contrast, the spectra score APV = 0.109 beyond R-ISS, age at diagnosis, and UAMS (**Table 3**) – also significant by likelihood ratio test (LRT, p = 7.4×10^-5^) – indicating that the spectra risk score contains predictive information beyond clinical factors and the previously established UAMS risk score.

**Table 3.** Overall survival Cox models including covariates and added predictive values.

| Variable | Base Model | $\Delta$ AIC | $\Delta$ BIC | APV | LRT *P* Value |
| --- | --- | --- | --- | --- | --- |
| UAMS | R-ISS + Age | -71.7 | -68.9 | 0.576 | $9.20 \times 10^{-18}$ |
| SBUK | R-ISS + Age | -52.7 | -49.9 | 0.502 | $1.42 \times 10^{-13}$ |
| OS Spectra | R-ISS + Age | -83.5 | -80.7 | 0.612 | $2.36 \times 10^{-20}$ |
| UAMS | R-ISS + Age + SBUK | -17.8 | -15.0 | 0.154 | $8.58 \times 10^{-6}$ |
| UAMS | R-ISS + Age + OS Spectra | -1.9 | 0.8 | 0.027 | 0.048 |
| SBUK | R-ISS + Age + UAMS | 1.2 | 4.0 | 0.006 | 0.371 |
| SBUK | R-ISS + Age + OS Spectra | 1.1 | 3.8 | 0.007 | 0.333 |
| OS Spectra | R-ISS + Age + UAMS | -13.7 | -10.9 | 0.109 | $7.39 \times 10^{-5}$ |
| OS Spectra | R-ISS + Age + SBUK | -29.7 | -27.0 | 0.226 | $1.77 \times 10^{-8}$ |
| OS Spectra | R-ISS + Age + UAMS + SBUK | -12.9 | -10.2 | 0.104 | $1.13 \times 10^{-4}$ |

R-ISS: Revised international staging system
Age at diagnosis
$\Delta$ AIC: Change in Akaike's Information Criterion by adding the variable
$\Delta$ BIC: Change in Bayesian information criterion by adding the variable
APV: Variable added predictive value of the variable
LRT *P* Value: Likelihood ratio test p-value between the base model with and without the variable

Eleven patients had at least three follow-up CD138+ samples, spanning up to six years. To illustrate a potential to track changes over time, the OS spectra risk score was graphed for the initial and longitudinal follow-up samples (**Figure 5**).

**Figure 5.**
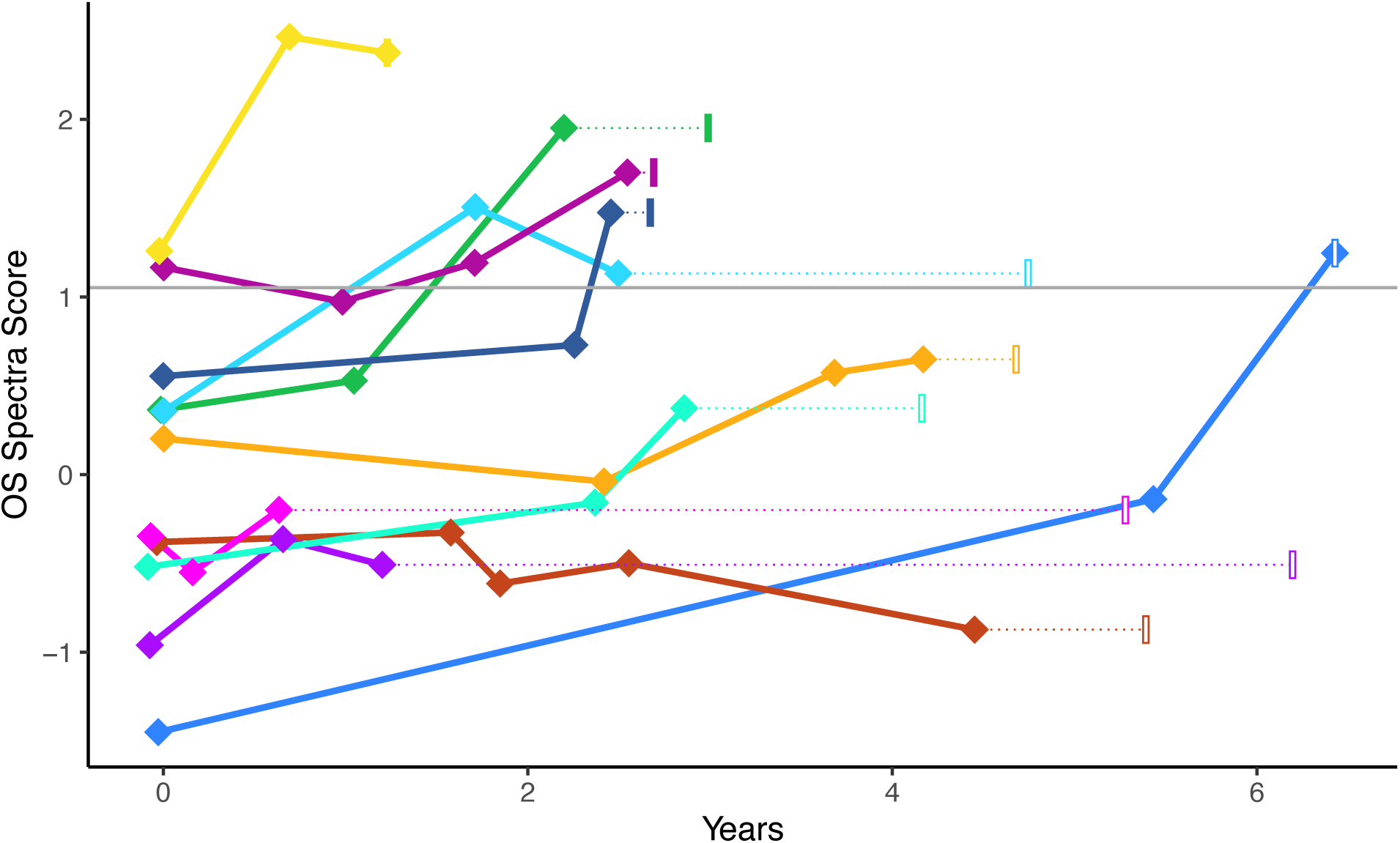
Aggregate effect of CD138+ spectra for overall survival over time. Spectra scores for overall survival are shown in eleven patients with RNAseq data at multiple time points. Diamonds indicate sequencing events and show the spectra score at that timepoint. The final narrow rectangle indicates timepoint the patient died (filled) or was last known alive (open). Gray horizontal line shows the high/low risk group threshold.

### Spectra Predictive Modeling – Progression-Free Survival (PFS)

As the spectra are unsupervised, the same 39 spectra variables can be used to model any outcome. We repeated the same predictive modeling procedure for PFS. Cox regression of PFS (392 events) selected nine spectra in a model with C_adj_ = 0.60 (0.60-0.67). Four of the retained spectra were distinct from the OS spectra model (**Table 1**). Gaussian mixture modeling identified two risk groups (p = 0.001) (**Figure S2**). Median time to progression was 9.7 months in the high-risk patients (n = 60, 50 events) and 35.7 months in the low-risk patients (n = 707, 342 events) (**Figure 6A**). Between high and low risk groups, spectra PFS HR_adj_ = 3.08 (1.67-8.44) while UAMS PFS HR_adj_ = 2.40 (1.70-3.31) and SBUK PFS HR_adj_ = 1.92 (1.59-2.43) (**Table S1**). Spectra added information beyond R-ISS, age at diagnosis, and UAMS (APV = 0.24, LRT p = 8.2×10^-8^) in predicting PFS (**Table S2**).

**Figure 6.**
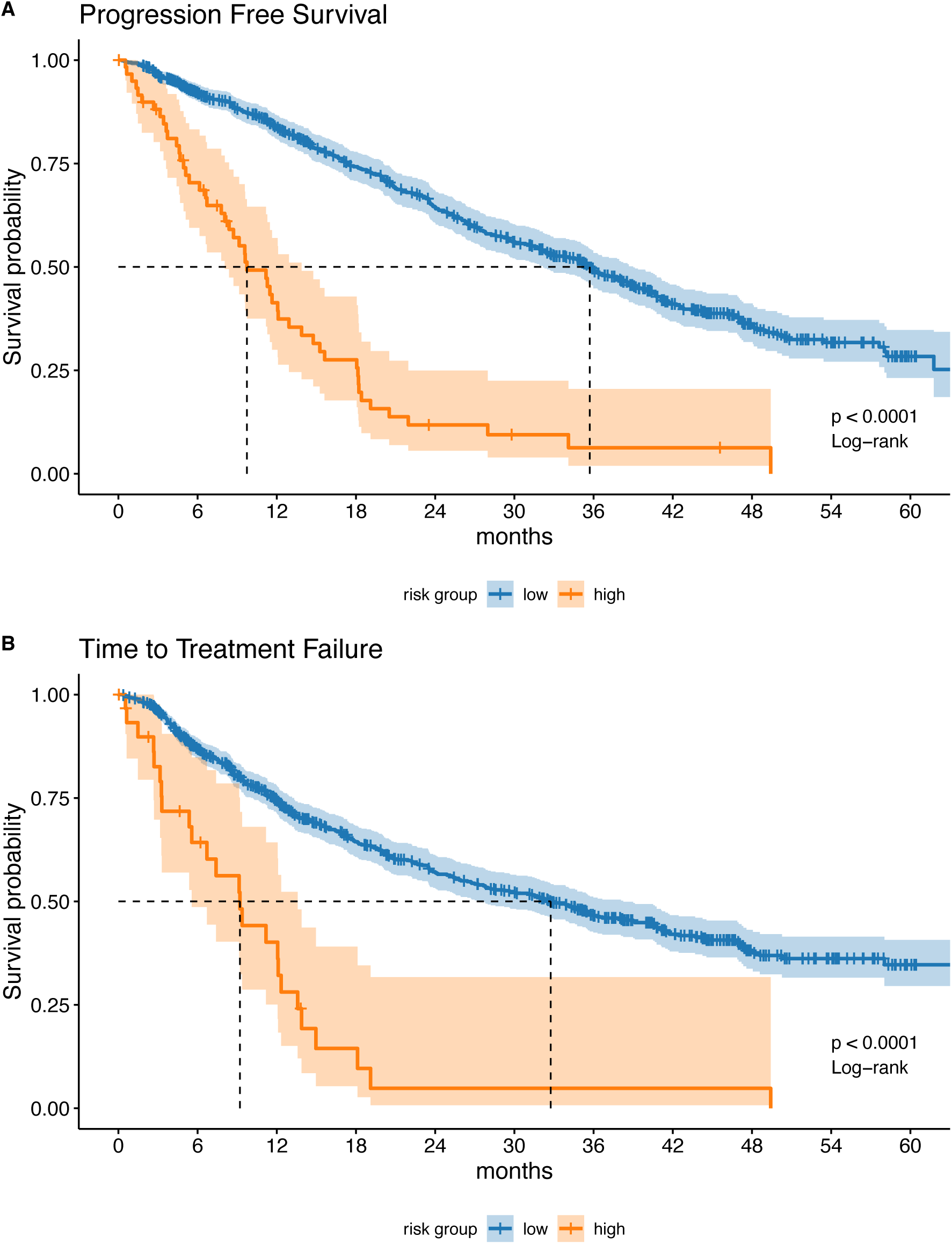
Kaplan-Meier curves of progression free survival and time to first line treatment failure. A) PFS high-risk patients (n = 60, 50 events) had median survival of 9.7 months. PFS low-risk patients (n = 707, 342 events) had median survival of 35.7 months. B) Patients in the spectra high-risk TTF (n = 31, 25 events) had median TTF of 9.2 months compared to low-risk patients (n = 736, 344 events) with median TTF of 32.8 months.

### Spectra Predictive Modeling – Time to First-Line Treatment Failure (TTF)

In Cox regression modeling to predict TTF (369 events) ten spectra were selected with the model C_adj_ = 0.60 (0.59-0.66). Four of the retained spectra were distinct from the OS and PFS models (**Table 1**). Two risk groups were identified in the TTF spectra score with GMM (p = 0.008) (**Figure S3**). Patients in the spectra high-risk TTF (n = 31, 25 events) had median TTF of 9.2 months compared to low-risk patients (n = 736, 344 events) with median TTF of 32.8 months (**Figure 6B**). Hazards ratios between high- and low-risk groups using TTF spectra risk groups (HR_adj_ = 3.10 [1.31-5.46]) outperformed UAMS (TTF HR_adj_ = 1.98 [1.48-2.68]) and SBUK (TTF HR_adj_ = 1.85 [1.52-2.36]) (**Table S3**). Spectra provided additional information in predicting TTF beyond R-ISS, age at diagnosis, and UAMS (APV = 0.29, LRT p = 2.6×10^-6^) (**Table S4**).

### Spectra Descriptive Modeling

**Figure 7** illustrates associations between the CD138+ spectra and tumor aberrations or demographic groups with elevated myeloma risk. For descriptive modeling a model containing all 39 spectra was fit for each characteristic. In logistic regression models, each tumor aberration showed different significant spectra in the model, with some spectra unique to only one aberration. Linear regression with age at diagnosis as a quantitative outcome highlighted associations with thirteen spectra. Seven spectra were significantly associated with gender and thirteen spectra significant with race (self-reported black or white; other racial categories too small to consider).

**Figure 7.**
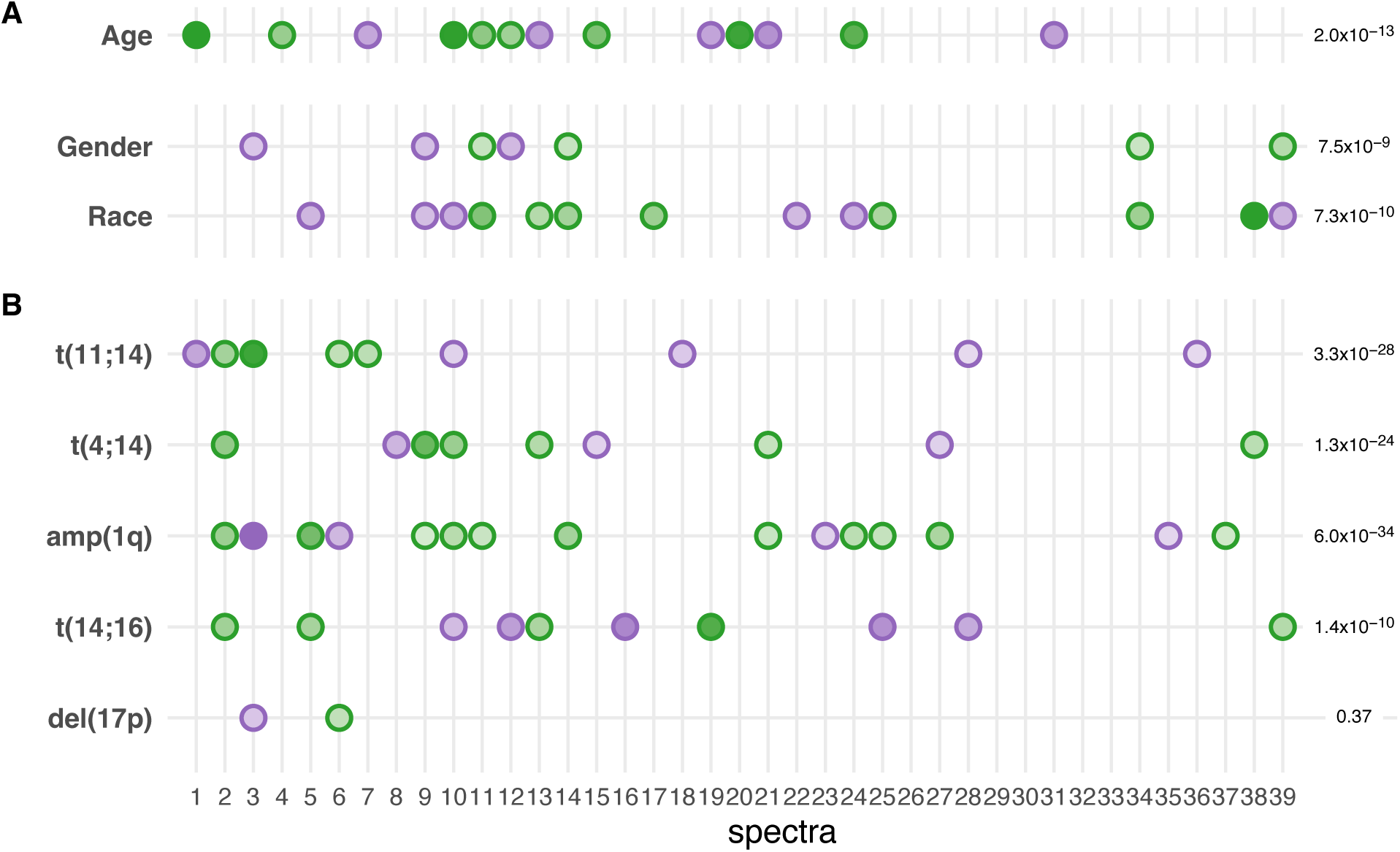
Overview of CD138+ spectra descriptive modeling. Multivariable modeling results for A) demographic characteristics and B) tumor FISH aberrations. The outcome of interest (left y-axis) and the significance of the full 39-spectra model (right y-axis). Spectra variables are illustrated on the x-axis. The effect size (intensity) and direction of each spectrum are indicated: green negative beta coefficient, purple positive beta coefficient. Effect size (intensity) is scaled to each plot (separated by a horizontal white space). No dot is shown if a spectrum was not significantly associated at *p* < 0.05 level.

## DISCUSSION

The promise of personalized prevention, management, and treatment is rooted in an ability to describe an individual’s unique experience and model important sources of heterogeneity (Ramón y Cajal et al., 2020). Gene expression in diseased tissue is an established source of heterogeneity (Kwa et al., 2017). Tools that can take a deeper dive and better characterize expression heterogeneity will be important to advance the promise of personalized medicine. Clinical and epidemiologic studies that wish to model multiple sources of risk in a population, transcriptome variables that can be easily incorporated with other variables and across endpoints of interest are advantageous. The goal of this study was to provide a technique to derive an agnostic framework of variables for transcriptome data, to empower multivariable studies, and provide novel quantitative molecular phenotypes. The SPECTRA approach identifies quantitative, orthogonal (non-correlated) variables that capture sources of transcriptome variation for use in subsequent predictive or descriptive modeling or as quantitative phenotypes.

The SPECTRA approach provides a measured dive into the transcriptome. Each spectrum iteratively moves quantifiably deeper into the variance of the data (measurable by its corresponding *λ*). Methods that iteratively find independent components (PCA and independent component analysis) have previously been shown to provide superior coverage of transcriptome data (Way et al., 2020). The importance of a deeper dive is illustrated in the MM predictive modeling. Spectra representing variance deeper in the data were important in predicting OS, PFS, and TTF (**Table 1**). For example, S32 (which explains 0.5% variance in the transcriptome) provided the same weight to the PFS risk score as S1 (8.7% variance). As another illustration, S19 and S26 (both with variance < 1%) are in the OS spectra risk score. Further, twelve patients with poor survival – 14% of the high-risk group – would not have been classified in the high-risk group if their S19 and S26 spectra values had been at the population mean. In our MM study, retention of components deep in the data, representing small variances (i.e., deep dives) improved prediction. We suggest that deep transcriptome characterization is a new tool with the potential to identify the few patients that respond to a drug, or small groups of individuals with large effects in outcome studies, both relevant to precision medicine. Furthermore, as shown previously for the PAM50 gene panel, superior coverage of data may identify previously overlooked expression differences between familial and sporadic tissues, identifying potential for familial components (Madsen et al., 2018), and providing new avenues for gene discovery, exposure and gene by environment studies.

In the SPECTRA approach negative gene loadings are embraced. Non-negative matrix factorization (NMF), arguably the leading approach in the computational biology field, restricts all values in the amplitude matrix (equivalent to the eigenvector matrix) and pattern matrix (equivalent to the transformed component values in PCA) to be non-negative. However, non-negative matrix values may not be a natural restriction to *systems of genes.* Genes in a system may act in opposite directions producing both surplus and deficits of that system. With the non-negativity restriction, NMF is limited to the identification of groups of over-expressed genes (Brunet et al., 2004; Stein-O’Brien et al., 2018), and thus models only neutrality and surplus. Deficits may also be important. While PCA spectra may represent mixtures of different biological mechanisms, these may be important combinations, including genes acting in opposite directions, and may better reflect reality. These differences underscore SPECTRA as a valuable and complementary tool to existing approaches.

In our MM study, we illustrated the implementation of 39 CD138+ spectra in predictive modeling for OS, PFS, and TTF. We showed added value beyond established risk scores (UAMS, SBUK) and clinical risk factors (R-ISS, age at diagnosis). The framework of 39 quantitative variables has the potential to provide a bridge to compare many patient or tissue characteristics (**Figure 7**) and could be used to compare existing categorizations (such as subtypes) of patients, even when no genes overlap in their signatures, or they predict different outcomes (Szalat et al., 2016). In this way, spectra provide an alternate to categorical intrinsic subtyping, a well-established practice for many cancers (Dai et al., 2015).

Clinically-relevant stratification may be better represented using risk groups within a transcriptome framework (Camp et al., 2019). To this end, we have described a rigorous strategy to define spectra-based risk-groups using GMM (**Figure 2**, **Figure 4**, **Table 2**). Using our approach to identify risk groups for OS, we classified 27 patients as high risk that UAMS classified as low risk (**Figure S4**). These 27 patients had median survival of 26.7 months. Conversely, the 35 patients classified as low risk by spectra score but high risk by UAMS did not reach median survival within the study timeframe.

SPECTRA outperform existing classification strategies in MM, while also allowing the flexibility of modeling with quantitative variables and multiple outcomes.

Descriptive modeling showed significant associations between spectra and tumor cytogenetics, race, gender, and age a diagnosis (**Figure 7**). As expected for a framework of agnostically derived variables, not all spectra are relevant to every dependent variable. Across the five tumor abnormalities, the number of associated spectra ranged from 2 to 15 with no single spectra associated with all abnormalities (**Figure 7**). Importantly, these examples show the flexibility of the framework as well as how it can support comparisons across different models and outcomes.

The potential for increased power using spectra variables is illustrated by the discovery of novel associations between spectra and patient demographic risk groups with known differences in incidence (age, gender, race). A prior CoMMpass study whose goal was to identify differences by race for myeloma tumors used the UAMS score to compare transcriptomes by race and did not identify significant differences (p = 0.662) (Manojlovic et al., 2017). In contrast, our framework of 39 CD138+ spectra did identify 13 spectra that differed significantly by race (**Figure 7**). Our multivariable results demonstrate that significant differences do exist, but also illustrate that the diseased cells in these demographic groups are not distinct entities; fewer than half the spectra variables differ significantly by these patient demographic groups. Focusing on the spectra that do show differences by demographics provides new avenues to explore why incidence varies in these groups, a key to disease prevention, intervention, and control. Because transcriptomes capture both the effects of internal (inherited genetics) and external factors (lifestyle, exposures, consequences of access to care), these results support epidemiology and biosociology investigations into such differences. We have provided the variable framework (gene transformations) and the spectra variables for the CoMMpass patients (see Data and Code Availability) to enable further study of spectra in other CoMMpass studies, as well as in other myeloma studies.

As for any approach, there are limitations. A key question is representation. For epidemiology studies, for example, spectra should ideally be representative of the entire disease population. This requires that the derivation dataset is a random sample from that population or based on a known selective sampling scheme. While there are many publicly available transcriptome datasets (Cancer Genome Atlas Research Network et al., 2013; GTEx Consortium, 2013), most fall short of this ideal. Thus, the spectra variables derived from these will have inherent limitations in representation. An investigator should consider if a derivation dataset is adequate to represent their study goals. We note that the goal of the MMRF CoMMpass study was designed to represent myeloma patients from diagnosis through treatment and is the largest existing cohort of treatment naïve CD138+ transcriptomes, with sampling continuing over time. To limit overfitting in spectra derivation, we used dimension reduction to focus only on the first *k* spectra (largest *k* components of variation), selected using a scree test (Cattell, 1966).

A limitation of agnostic variables is interpretation. To illustrate the ability to gain biological insight from spectra, we implemented pre-ranked GSEA (Subramanian et al., 2005). Each spectra variable is a linear transformation based on gene weights from its eigenvector. Gene weights can be positive or negative and order the importance and direction of effect of genes in each spectrum, and thus are an ideal ranking metric for enrichment. Spectrum variable S5 was significant in models for OS, PFS and TTF. We used fast-GSEA and identified enriched Hallmark pathways based on genes ranked by CD138+ S5 (Korotkevich et al., 2016; Liberzon et al., 2015). Three pathways were highly significant. Of particular interest was the unfolded protein response (UPR) pathway (normalized enrichment score, NES = 2.25, adjusted-p=2.5×10^-8^). Secreted proteins are processed in the endoplasmic reticulum (ER). Incorrectly folded proteins create ER stress and activate the UPR pathway, targeting them for degradation by the proteasome. Bortezomib is a proteasome inhibitor and hence causes build-up of misfolded proteins, increasing ER stress leading to cell-death. Myeloma cells secrete large amounts of incorrectly folded immunoglobulins and therefore are near capacity for UPR. Due to this, CD138+ myeloma cells are particularly sensitive to proteasome inhibitors, such as Bortezomib, a common agent in the initial treatment for MM. Thus, spectrum S5 may represent a patient’s sensitivity to proteasome inhibition agents, influencing OS, PFS and TTF.

Data quality and processing are paramount to derive informative agnostic variables. PCA is a procedure that provides linear transformations of the data to represent variance. If the data have technical artifacts, batch effects, unstable or non-comparable expression measures, the noise can overwhelm authentic variance. Accordingly, the SPECTRA approach intentionally includes strict quality control, careful zero-handling, robust normalization, and batch correction (**Figure 2**). Without these steps, PCA can fail to provide informative variables. An agnostic approach permits stringent data culling because the incentive to retain features based on known functional relevance is removed. The impetus is to only retain features that can contribute to meaningful variance and provide informative variables for modeling. The limitation of an agnostic approach is reduced biological interpretation or mechanistic insight of the variables themselves, prior to modeling. However, there are already many approaches that take this alternate goal of intermediate interpretation (Subramanian et al., 2005), whose limitations are instead the reduced flexibility of the variables they produce. Hence, SPECTRA is a complementary approach to the current toolset available for all fields.

In conclusion, we present a new approach, SPECTRA, to derive an agnostic transcriptome framework of quantitative, orthogonal variables for a dataset. These multi-gene expression variables are designed specifically to capture transcriptome variation, providing variables for flexible modeling, along with other covariates, to better differentiate individuals for any outcome. Spectra may also be used as novel transcriptome phenotypes. Applied to CD138+ transcriptomes for myeloma patients, we defined CD138+ spectra and implemented these in many different outcome models. We illustrated an ability to predict prognostic outcomes and provide new insight into potential differences between tumors and patients from demographic groups. Fundamentally, the technique shifts from characterizing a transcriptome using categories to multiple quantitative measures. The SPECTRA approach provides a new paradigm and tool for exploring transcriptomes that hold promise for discoveries to advance precision screening, prevention, intervention, and survival studies.

## MATERIALS AND METHODS

### SPECTRA approach

Our goal was derivation of well-behaved, quantitative variables from RNAseq data to capture the many sources of intrinsic variation in a transcriptome. To represent meaningful variance and enable deep comparison across individuals we model well-behaved genes after normalization and batch correction (Figure 2).

Features in the transcriptome likely to be unduly influenced by poor alignment or lacking precision due to sequencing depth are removed as potentials for introducing spurious and unstable variation. We concentrate on autosomal protein-coding genes. Features with inadequate data for precision, defined as more than 5% of samples with fewer than 100 read counts, are removed. We also remove genes known to be unstable across different RNAseq pipelines (Arora et al., 2020). After the removal of features, individuals are removed from consideration if more than 10% of the remaining genes have fewer than 100 read counts.

Normalization is required for comparisons across genes and individuals and includes adjustment for gene length, sequencing depth (library size), and RNA composition. We use a robust internal (single sample) normalization to obviate the need for a ‘reference’ sample and to provide the possibility for portability across datasets. The SPECTRA approach is gene-focused, but the workflow handles both gene-based and transcript-based alignment and quantification; the latter has been suggested to be more accurate (Zhao et al., 2015). Normalized gene expression values, e*_g_*, are calculated as follows:

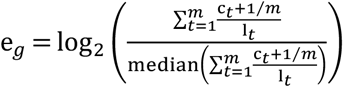

where *c_t_* is the read count for transcript *t*, l*_t_* is the transcript length in kilobases (extracted from the GTF used to align and quantify the RNAseq data), and *m* is the number of transcripts for the gene. Zero-handling is achieved by adding 1/*m* to the transcript counts (c*_t_* + 1⁄*m*). Division by l*_t_* corrects for transcript length. Summing the length-corrected transcript counts results in a gene-level count per kilobase (CPK) measure. Gene-level read counts are equivalent to *m* =1. Adjustments for sequencing depth and RNA composition (often referred to as the *size factor*) are achieved via the division of each gene-based CPK measure by the median of CPK-values for retained features. We note that the more usual upper-quartile adjustment also provides robust internal normalization (Shahriyari, 2019); however, since our implementation is post-QC after numerous features have been removed for low counts, the median is more suitable. Normalized data are *log_2_* transformed to account for skew. We also truncate outliers to a five standard deviation threshold from the mean of the normalized gene counts in the relevant direction.

Batch correction is necessary to correct for potential technical artifacts and spurious variation introduced by differing sequencing protocols. We adjust for sequence batch using ComBat (Johnson et al., 2007) as implemented in the SVA R package (Leek et al., 2012), with covariates included for patient characteristics that are unbalanced by batch.

PCA is implemented with the covariance matrix. We use singular value decomposition to perform PCA, and it is necessary to center the expression values first to ensure the MF is performed for the covariance. Expression values (*e_g_*) are centered on the mean across individuals for gene *g*. The R core function *prcomp* is used to perform PCA. Eigenvectors contain the gene loadings that define the linear transformation used for each spectrum. A corresponding eigenvalue, *λ*, indicates the proportion of the global variance represented by the transformed value for an eigenvector. We use a scree test(Cattell, 1966) (inflection point of the rank-ordered plot of the *λ*, or elbow method) to select the *k* components to retain. The proportion of variance explained by this *k*-dimensional space 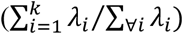 indicates the depth of the dive into the transcriptome data. Spectra variables are the standardized retained components (S_1_, …, S*_k_*).

### Myeloma CD138+ Spectra

Data from the MMRF CoMMpass Study (release IA14) (Keats et al., 2013) were downloaded from the MMRF web portal (https://research.themmrf.org/). Clinical data and CD138+ RNAseq were available for 768 patients prior to treatment at study entry (baseline) and 119 follow-up bone marrow samples. Transcript-based expression estimates processed by Salmon (version 0.7.2) were used. The SPECTRA approach was used on baseline samples (n=768) to derive a CD138+ transcriptome framework and SPECTRA variables. While not used for spectra derivation, follow-up samples were batch-corrected alongside the baseline samples. Covariates included in batch correction were age, gender, overall survival, progression-free survival, time to first-line treatment failure, and treatment status. The first 39 components (spectra variables S1, …, S39). were selected based on the scree test (Cattell, 1966).

### Predictive Modeling of Clinical Outcomes

Predictive Modeling used Cox proportional hazards regression, implemented in the *survival* package in R (Therneau & Grambsch, 2000; Therneau, 2021). An overview of predictive modeling is provided in **Figure S1**. All 39 spectra were considered as predictor variables for each of the three survival outcomes: overall survival (OS), progression-free survival (PFS), and time to first-line treatment failure (TTF). The spectra were standardized in all analyses. For each outcome, a single step variable selection from the all-spectra model was performed to retain only significant spectra (p<0.05). The linear predictor from the reduced model was used to define a quantitative spectra risk score for the outcome (i.e., the weighted sum of the spectra retained in the model based on their coefficients: ∑*_j_ β_j_S_j_*).

To determine if a quantitative spectra risk score displayed evidence of latent risk groups that could be clinically meaningful, the risk score distribution and normal Quantile-Quantile plots were inspected. The *mclust* R package was used to statistically assess evidence for risk groups using density estimation by Gaussian finite Mixture Modeling (GMM) (Fraley & Raftery, 2002; Scrucca et al., 2016) assuming equal variances. Bayesian information content (BIC) and bootstrap likelihood ratio tests (LRT) were used to determine the number of risk groups (*mclustBIC, mclustBootstrapLRT*). The hazard ratio (HR) between risk groups was calculated using relative event rates (Armitage et al., 2002, p. 578) from *survdiff* in the *survival* package in R (Therneau, 2021).

To address overfitting and lack of a replication sample, bootstrap internal validation was used (Harrell, 2015, pp. 114–116). Bootstrap internal validation involves replicating the entire modeling process on bootstrap resamples of the data to determine and adjust for over-fitting. Here, the process includes both the single step variable selection, GMM density estimation for risk group designation, and adjustment of HRs, C-index and bootstrap confidence limits (Noma et al., 2021). Initial estimates from a model are referred to as ‘apparent’. The measure of overfitting is referred to as ‘optimism’. The corrected estimates are referred to as ‘adjusted’ values. Adjusted estimates are more reasonable assessment of effect size. If adjusted confidence intervals do not contain 1.0, this indicates validation.

#### Comparison with existing transcriptome risk scores

We compared spectra-based risk to two previously published risk scores: 1) the first and most widely adopted supervised expression risk score in myeloma, from the University of Arkansas for Medical Sciences (UAMS) (Shaughnessy et al., 2007); and 2) a recent supervised risk score, from the Shahid Bahonar University of Kerman (SBUK) (Zamani-Ahmadmahmudi et al., 2020). The UAMS risk score was developed in microarray data and tested 54,657 probes for association with disease-related survival (Shaughnessy et al., 2007). A total of 70 genes were selected (19 under and 51 overexpressed prognostic genes). The UAMS risk score is the ratio of mean expression of the up-regulated to down-regulated genes, with k-means clustering to determine a cutoff for ‘high-risk’ classification (Shaughnessy et al., 2007). The SBUK prognosis score was developed in the CoMMpass RNAseq data. All genes were tested for association with survival, followed by a multi-step process, including univariate Cox analysis, the intersection in six Class Prediction algorithms, and multivariate Cox analysis, was used to select 17 genes consistent across multiple methods (Zamani-Ahmadmahmudi et al., 2020). These 17 were entered in a multivariable Cox regression to define the SBUK score, with the 75^th^ percentile used to classify patients into high-risk and low-risk categories (Zamani-Ahmadmahmudi et al., 2020). We calculated each patient’s UAMS and SBUK risk scores and their risk status (low or high) using the CoMMpass RNAseq data.

#### Added predictive value

Predictive value beyond established clinical risk factors is important to establish. We calculated the added predictive value (APV) (Al-Radi et al., 2007; Califf et al., 1985; Harrell, 2015) of each expression risk score (spectra, UAMS, SBUK) beyond risk predicted from established clinical predictors. We used quantitative risk scores, and not risk group status, as these contain more complete risk information and because each gene-expression method employed a different thresholding approach making risk status comparisons potentially misleading. The APV compares the LRT statistic for a model including established clinical predictors to a model which also includes each expression risk score. We also determined whether the spectra risk score provided value beyond known clinical predictors and UAMS. APV=0.0 indicates no additional prediction information, i.e., all predictive power is duplicative with other factors already in the model. With larger independent predictive contributions, APV increases, to a maximum of 1.0.

### CD138+ Spectra in Follow-Up Samples

To illustrate the potential to track changes over time, OS spectra risk scores were calculated in longitudinal follow-up samples. For these samples, normalized and batch-corrected gene expression values were centered to the derivation set. Spectra values (S*_j_*) were determined using the previously derived linear transformation and scaled to the derivation set. The OS risk score was calculated using the previously established linear combination across spectra (∑*_j_ β_j_S_j_*). Spectra variables in all samples (baseline and follow-up) are provided (see Data and Code Availability).

### Descriptive Modeling of Clinical and Demographic factors

We used descriptive modeling to illustrate associations between the CD138+ spectra and clinical and demographic factors with elevated myeloma risk: tumor aberrations (high risk del(17p), t(14;16), gain 1q, and t(4;14), and standard risk t(11;14)) (Rajkumar & Kumar, 2020), age, gender, race. Linear or logistic regression was used with all 39 CD138+ spectra modeled as independent variables. Spectra were noted if significant in the descriptive model (p < 0.05).

### Data and Code Availability

Processed RNAseq data from the CoMMpass Study can be downloaded from https://research.themmrf.org/. Code used to derive the CD138+ transcriptome spectra and generate the myeloma results is freely available on GitHub: https://github.com/njcamp-lab/MM_spectra. Details of the QC process and the transcriptome framework (linear equations for the gene transformations) necessary to calculate the 39-spectra variables in other studies are also provided. Spectra for individuals in the IA14 CoMMpass data can be downloaded from https://github.com/njcamp-lab/MM_spectra/SpectraData.

## Data Availability

All data is publicly available through the MMRF Researcher Gateway (https://research.themmrf.org).

https://portal.gdc.cancer.gov/projects/MMRF-COMMPASS

## ACKNOWLEDGMENTS

The research reported in this publication was supported by the National Cancer Institute (Award Numbers F99CA234943, K00CA234943, K07CA230150, and P30CA042014-29S9), the National Center for Advancing Translational Sciences (Award Number UL1TR002538), and the National Library of Medicine (Award Number T15LM007124) of the National Institutes of Health. The content is solely the responsibility of the authors and does not necessarily represent the official views of the National Institutes of Health.

## COMPETING INTERESTS

None.

## SUPPLEMENTAL METHODS

Here we establish the matrix factorization (**MF**) natural for individual-based outcome modeling. Data matrices, ***X*** and 𝑻, are *oriented with individuals as subjects (n rows) and genes as variables (g columns)*. Given a *n* × *g* design matrix, ***X*** (mean-centered expression values for *n* individuals on *g* genes), PCA is the MF

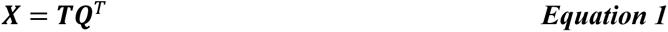

where ***T*** contains the transformed values (the dimension variables), and ***Q*** is the PCA ‘rotation’ matrix. Each row in ***Q****^T^* = (***q***_1_, ***q***_2_, … , ***q****_g_*)*^T^* is an orthogonal eigenvector (or component) which holds the coefficients for the linear model to transform the observed gene values into the spectra variables. The set of linear transformations are the transcriptome framework. The rotation matrix can be derived from the eigen decomposition of the covariance matrix, 𝚺

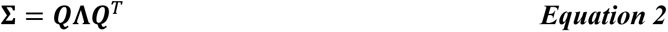

where 𝚺 is proportional to 𝑿*^T^* 𝑿, and 𝚲 is the diagonal matrix of eigenvalues. Each eigenvalue, *λ_S_*, is a scalar indicating the proportion of the global variance represented by the transformed value defined by the *s*^th^ eigenvector, ***q****_s_*, in 𝑸. Eigenvalues are ranked, such that the first PC, defined by ***q***_1_ captures the most variance, ***q***_2_ the next highest, and so on. We note that there can only be min(*n*, *g*) non-zero eigenvalues, because, beyond this no variance remains. In most, if not all, existing RNAseq studies, there are more genes than individuals and hence *n* is the limiting rank.

Dimensionality can be reduced to *k* dimensions by utilizing ***Q****_k_*; only the first *k* columns (PCs) of ***Q***. After selection of *k* PCs, transformed values are represented as:

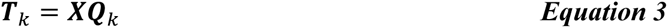

We note that PCA is deterministic and therefore the selection of *k* is a post-procedure decision that does not influence the MF. The proportion of variance explained by the retained dimensions 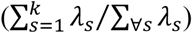 can be used as a measure of coverage.

## SUPPLEMENTAL TABLES

**Table S1.** Hazard ratios of high vs low risk of progression free survival.

| **Risk Score** | **Apparent HR** | **Adjusted HR** | **Optimism** |
| --- | --- | --- | --- |
| PFS Spectra | 4.22 (2.81-9.59) | 3.08 (1.67-8.44) | 1.14 |
| UAMS | 2.44 (1.74-3.35) | 2.40 (1.70-3.31) | 0.04 |
| SBUK | 1.94 (1.61-2.45) | 1.92 (1.59-2.43) | 0.01 |

**Table S2.**
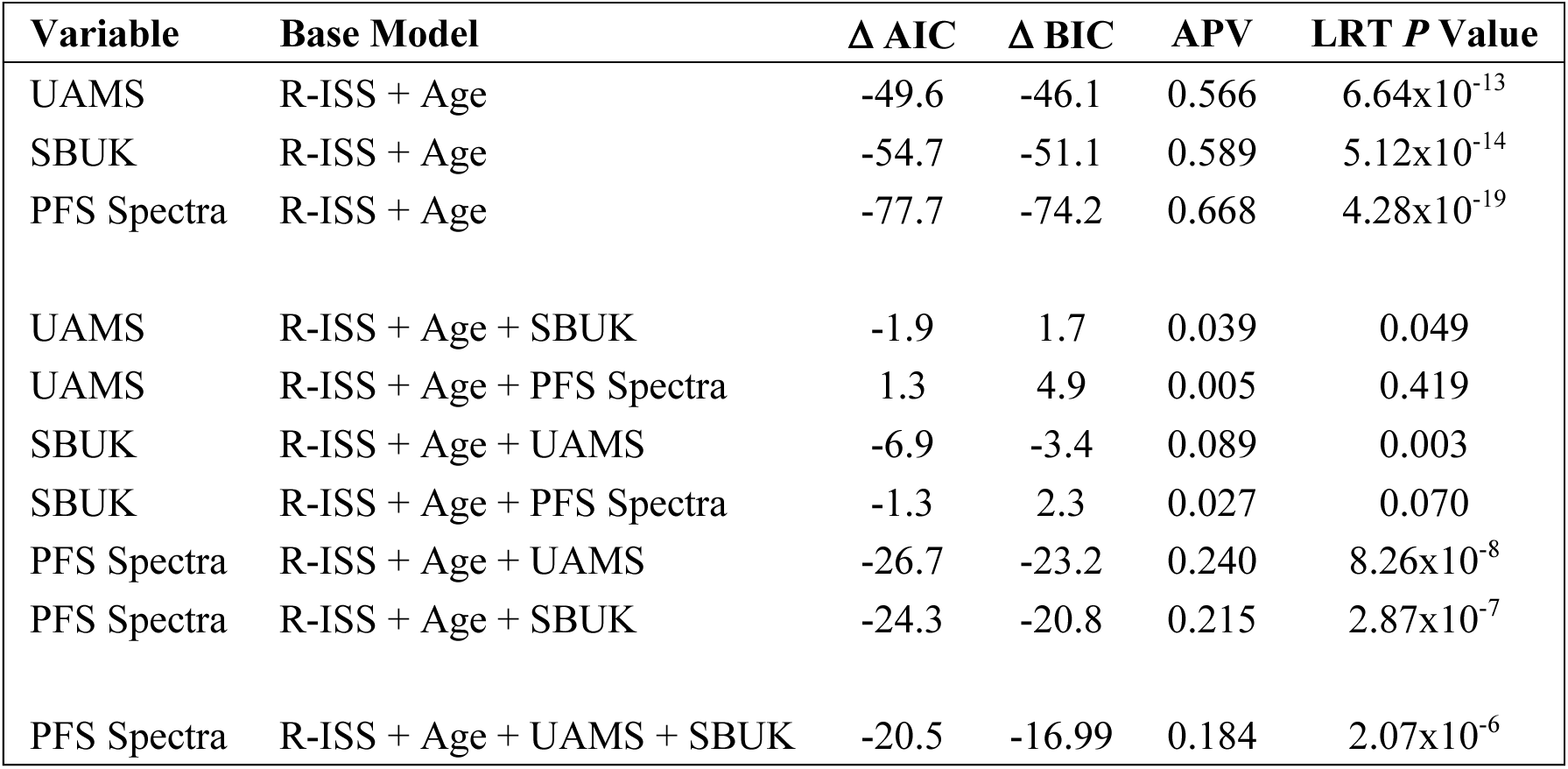
Progression free survival covariates and added predictive values.

| **Variable** | **Base Model** | **$\Delta$ AIC** | **$\Delta$ BIC** | **APV** | **LRT *P* Value** |
| --- | --- | --- | --- | --- | --- |
| UAMS | R-ISS + Age | -49.6 | -46.1 | 0.566 | $6.64 \times 10^{-13}$ |
| SBUK | R-ISS + Age | -54.7 | -51.1 | 0.589 | $5.12 \times 10^{-14}$ |
| PFS Spectra | R-ISS + Age | -77.7 | -74.2 | 0.668 | $4.28 \times 10^{-19}$ |
| UAMS | R-ISS + Age + SBUK | -1.9 | 1.7 | 0.039 | 0.049 |
| UAMS | R-ISS + Age + PFS Spectra | 1.3 | 4.9 | 0.005 | 0.419 |
| SBUK | R-ISS + Age + UAMS | -6.9 | -3.4 | 0.089 | 0.003 |
| SBUK | R-ISS + Age + PFS Spectra | -1.3 | 2.3 | 0.027 | 0.070 |
| PFS Spectra | R-ISS + Age + UAMS | -26.7 | -23.2 | 0.240 | $8.26 \times 10^{-8}$ |
| PFS Spectra | R-ISS + Age + SBUK | -24.3 | -20.8 | 0.215 | $2.87 \times 10^{-7}$ |
| PFS Spectra | R-ISS + Age + UAMS + SBUK | -20.5 | -16.99 | 0.184 | $2.07 \times 10^{-6}$ |

**Table S3.** Hazard ratios of high vs low risk early treatment failure.

| **Risk Score** | **Apparent HR** | **Adjusted HR** | **Optimism** |
| --- | --- | --- | --- |
| TTF Spectra | 3.93 (2.13-6.28) | 3.10 (1.31-5.46) | 0.82 |
| UAMS | 2.00 (1.50-2.70) | 1.98 (1.48-2.68) | 0.02 |
| SBUK | 1.86 (1.52-2.36) | 1.85 (1.52-2.36) | 0.01 |

**Table S4.** Time to first-line treatment failure covariates and added predictive values.

| **Variable** | **Base Model** | **$\Delta$ AIC** | **$\Delta$ BIC** | **APV** | **LRT *P* Value** |
| --- | --- | --- | --- | --- | --- |
| UAMS | R-ISS + Age | -33.5 | -30.1 | 0.478 | $2.49 \times 10^{-9}$ |
| SBUK | R-ISS + Age | -40.6 | -37.1 | 0.523 | $6.73 \times 10^{-11}$ |
| TTF Spectra | R-ISS + Age | -55.1 | -51.6 | 0.595 | $4.10 \times 10^{-14}$ |
| UAMS | R-ISS + Age + SBUK | 0.6 | 4.1 | 0.017 | 0.240 |
| UAMS | R-ISS + Age + TTF Spectra | 1.5 | 5.0 | 0.005 | 0.474 |
| SBUK | R-ISS + Age + UAMS | -6.4 | -2.9 | 0.102 | 0.004 |
| SBUK | R-ISS + Age + TTF Spectra | -0.4 | 3.1 | 0.024 | 0.124 |
| TTF Spectra | R-ISS + Age + UAMS | -20.1 | -16.6 | 0.229 | $2.61 \times 10^{-6}$ |
| TTF Spectra | R-ISS + Age + SBUK | -14.9 | -11.4 | 0.172 | $3.96 \times 10^{-5}$ |
| TTF Spectra | R-ISS + Age + UAMS + SBUK | -13.6 | -10.1 | 0.158 | $7.98 \times 10^{-5}$ |

## SUPPLEMENTAL FIGURES

**Figure S1.**
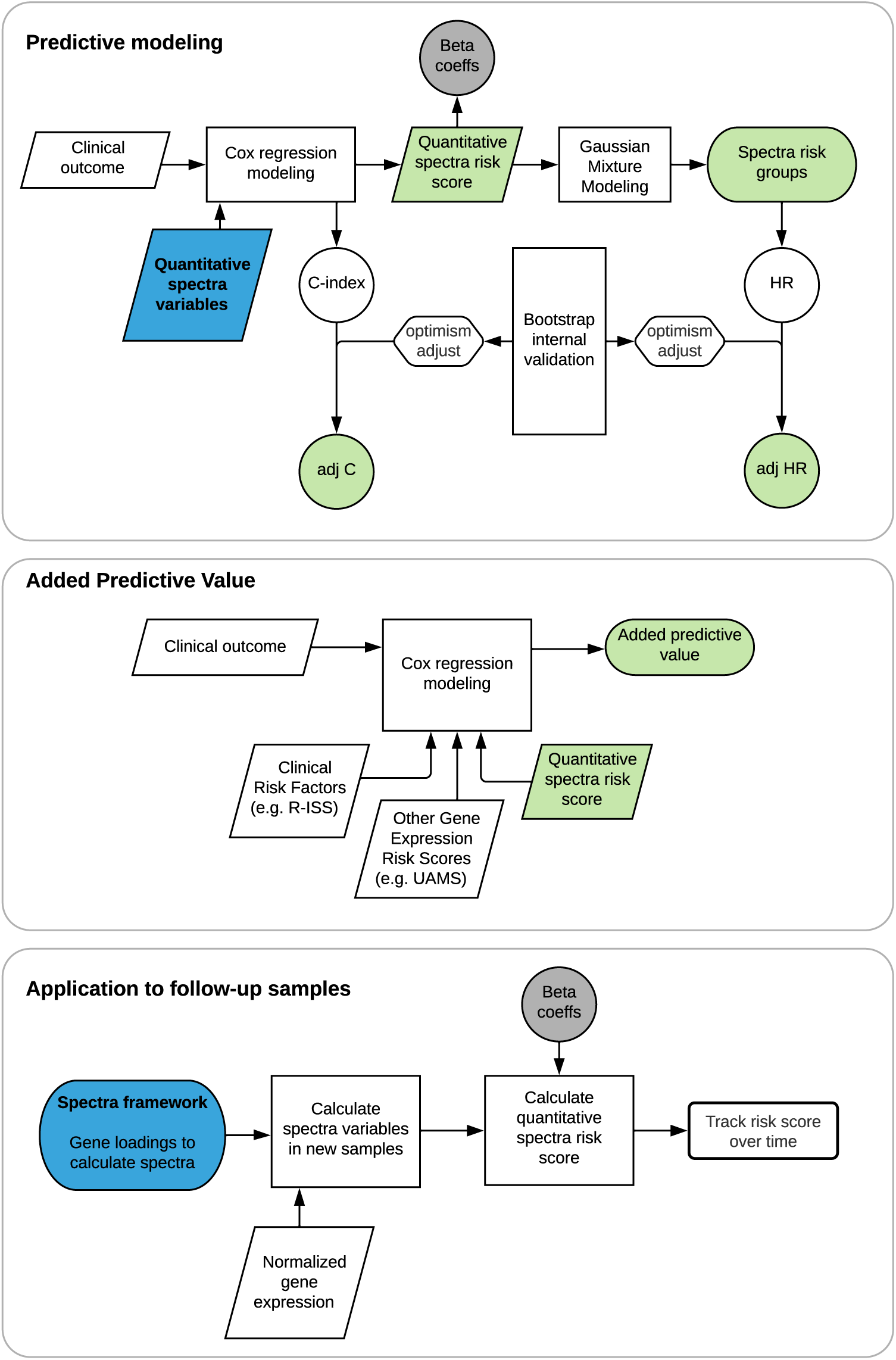
Diagram of predictive modeling.

**Figure S2.**
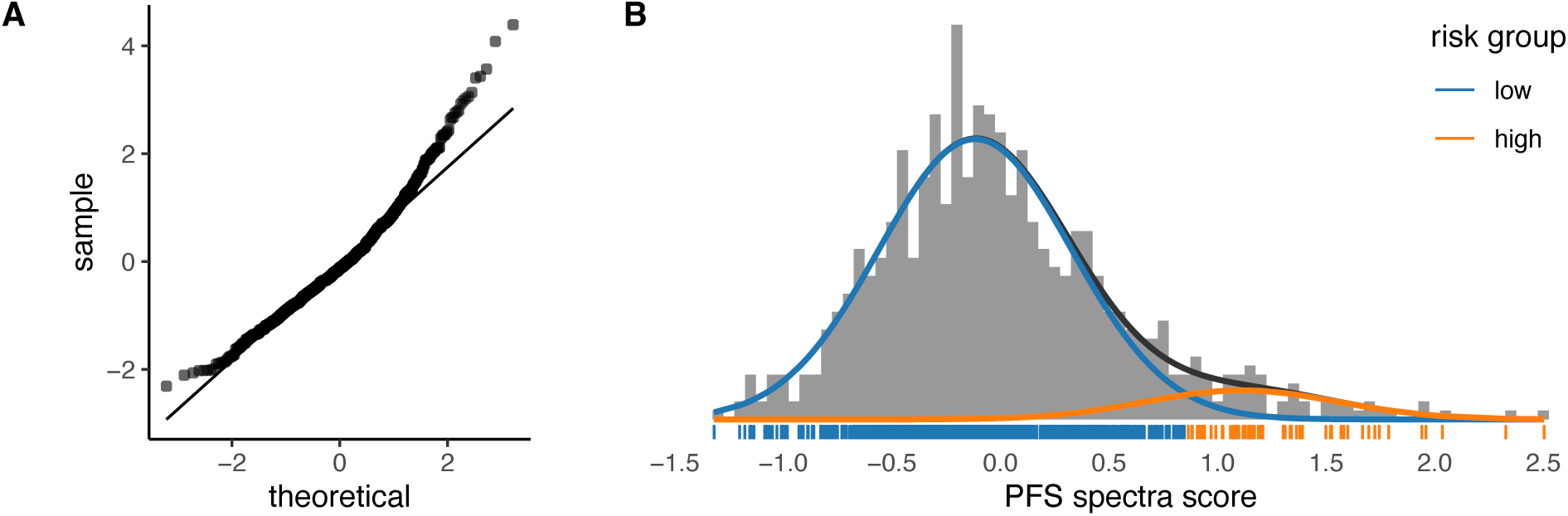
Progression free survival spectra score. A) Quantile-quantile plot. B) Gaussian mixture modeling to identify patients at high and low risk of progression.

**Figure S3.**
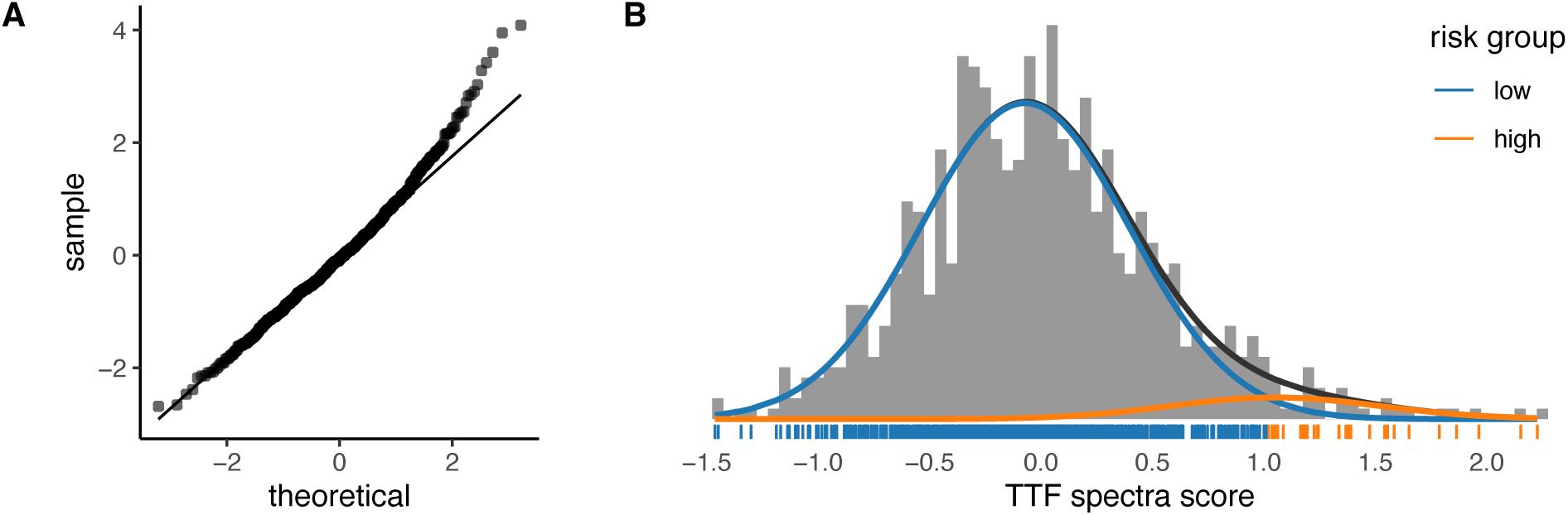
Time to first line treatment failure spectra score. A) Quantile-quantile plot. B) Gaussian mixture modeling to identify patients at high and low risk of early treatment failure.

**Figure S4.**
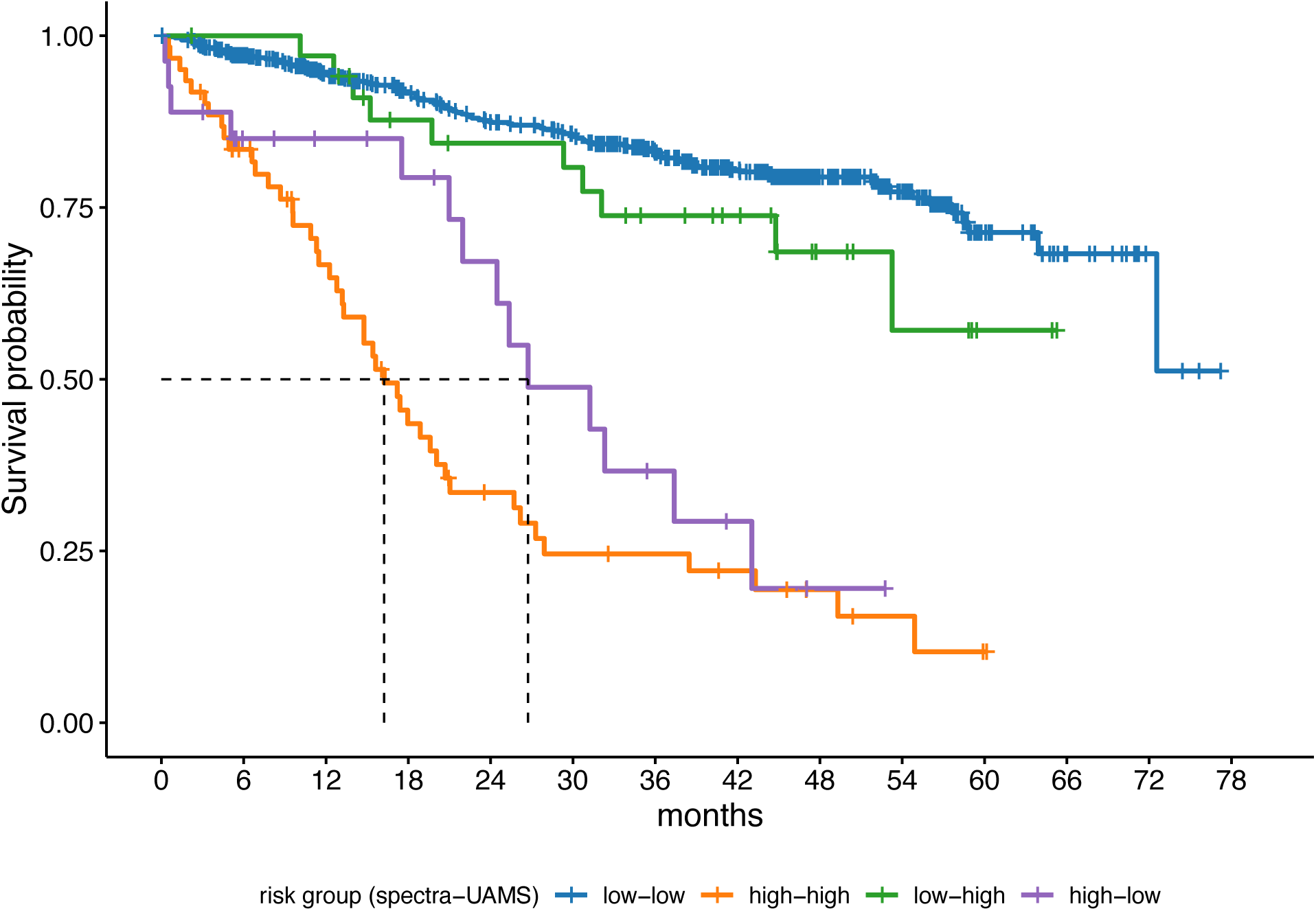
Kaplan-Meier curves of overall survival by spectra and UAMS risk-groups. Spectra identified patients as high-risk with worse survival that UAMS classified as low risk.

## Notes

### Competing Interest Statement

The authors have declared no competing interest.

### Author Declarations

Publicly available RNAseq data were used in this project from the Multiple Myeloma Research Foundation (MMRF) CoMMpass trial (NCT145429). To be eligible, a patient must have read, understood and signed informed consent. RNAseq data was downloaded directly from the MMRF Researcher Gateway (https://research.themmrf.org) after creating an account. Permission was also obtained to access these data via dbGaP under accessions phs000348.v2.p1 and phs000748.v4.p3.

